# The association between visual hallucinations and secondary psychosis: A systematic review and meta-analysis

**DOI:** 10.1101/2022.06.07.22276049

**Authors:** Amber Kaur Dadwal, Graham Blackman, Maria Teixeira-Dias, Dominic ffytche

## Abstract

**Background:** Visual hallucinations are often considered by clinicians to be suggestive of a secondary cause of psychosis, however, this association has never been assessed meta-analytically. We aimed to compare the presence of visual hallucinations in patients with psychosis due to a primary or secondary cause.

**Method:** A meta-analysis of case-control studies directly comparing primary and secondary psychosis was conducted. PubMed, OVID, MEDLINE, Embase, PsychINFO and Global Health databases were searched without date restrictions. Two researchers performed the search independently. The inclusion criteria were studies that reported the frequency of visual hallucinations in patients with primary and secondary psychosis and were published in English. A random-effects model, following the DerSimonian and Laird method, was used to pool studies and generate overall odds ratios (OR) and 95% confidence intervals (CI).

**Results:** Fourteen studies (904 primary and 804 secondary psychosis patients) were included. Visual hallucinations were significantly associated with secondary psychosis (OR = 3.0, 95% CI = 1.7-5.1, *p*<0.001) with moderate between-study heterogeneity (I^2^=70%). Subgroup analysis by type of secondary psychosis (organic, drug-induced, mixed) was non-significant. Analysis of the content of visual hallucinations (51 primary and 142 secondary psychosis patients) found hallucinations of inanimate objects more likely to be associated with secondary psychosis (OR = 0.1, 95% CI = 0.01-0.8, *p*=0.03).

**Conclusions:** Visual hallucinations were found to be strongly associated with a secondary cause of psychosis. The presence of visual hallucinations in a patient presenting with psychosis may serve as a potential ‘red flag’ for a secondary cause and warrant further investigation.

**Key messages:** *What is already known:* Psychotic symptoms may arise due to either an idiopathic primary (‘non-organic’) disorder or a secondary cause, such as an underlying medical (‘organic’) disorder. Whilst it is widely accepted amongst clinicians that visual hallucinations are associated with a secondary psychosis, this has never been established meta analytically.

*What this study adds:* Patients with a secondary cause of psychosis are approximately three times more likely to experience visual hallucinations than patients with a primary psychiatric disorder. The content of visual hallucinations in patients with a secondary cause also differs compared to patient with a primary cause.

*How this study might affect practice:* Findings provide the strongest evidence to date for routinely assessing patients for the presence of visual hallucinations as part of the standard psychiatric assessment. A positive finding should be considered a ‘red flag’ for a secondary cause.

## INTRODUCTION

The lifetime risk of developing a psychotic disorder is estimated to be 3%(1). In most cases, this is an idiopathic primary (‘non-organic’) psychotic disorder such as schizophrenia, however in some cases, patients may develop psychotic symptoms that are attributable to a secondary cause, such as an underlying medical (‘organic’) disorder, or a drug-induced state(2). Preliminary evidence suggests that patients with a secondary psychosis may be distinguishable based on psychopathological and phenomenological characteristics(3).

It is widely accepted amongst clinicians that visual hallucinations are associated with a secondary psychosis, with empirical evidence supporting this assertion(4–6). Furthermore, in patients experiencing visual hallucinations, the content of the visual hallucinations may also differ between those with a primary or secondary cause(4,6–8). However, whether visual hallucinations are more common in secondary psychosis has never been assessed meta-analytically. The primary aim of this meta-analysis was to determine whether visual hallucinations were associated with secondary causes of psychosis by pooling studies which compared the frequency of visual hallucinations in primary psychotic disorders with secondary causes. The secondary aim was to determine whether there were phenomenological differences in the content of visual hallucinations between patients with a primary or secondary aetiology.

## METHODS

We performed a meta-analysis of case-control studies comparing the presence of visual hallucinations in primary and secondary causes of psychosis. The study was registered with the open science framework (OSF; https://osf.io/j9twg/?view_only=5c0f63cccd574239aca07bccdae35f3c) and the Preferred Reporting Items for Systematic reviews and Meta-Analyses (PRISMA) guidelines were followed(9) (see table 1 in supplementary materials).

**Table 1.** Frequency and odds ratios between types of visual hallucinations experienced in patients with organic and non-organic psychosis. OR = odds ratios, CI = confidence interval. P-value based on χ^2^

| Visual hallucination | N (%) |  | OR | CI | z | p |
| --- | --- | --- | --- | --- | --- | --- |
|  | Primary | Secondary |  |  |  |  |
| Simple | 0 | 9 | 7.3 | 0.4-128 | 1.36 | 0.17 |
| Complex | 51 | 133 |  |  |  |  |
| Animate | 50 | 119 | 0.1 | 0.01-0.8 | 2.2 | 0.03* |
| Inanimate | 1 | 23 |  |  |  |  |

### Search strategy

The databases PubMed, OVID, MEDLINE, Embase, PsychINFO and Global Health were searched without date restrictions up to the 30^th^ October 2020. The following search terms were used: “Psychos*” AND “Schizophreni*” AND “Visual hallucinat*”. We restricted the search to studies published in English. The title and abstract of all studies identified were screened; thereafter full-text reviews were carried out to confirm eligibility. In addition, references of included studies were manually searched. The search was performed independently by two researchers (A.K.D and M.T.D) with discrepancies resolved by a third researcher (G.B).

### Eligibility criteria

Inclusion criteria were: i) a case-control study design where primary and secondary causes of psychosis were compared directly, ii) ascertainment of the presence or absence of visual hallucinations and iii) classification of patients into having a primary (i.e. non-organic) or secondary psychosis. The latter included both medical (‘organic’) causes, as well as drug-induced states.

### Extracted variables

The primary variable extracted was the number of patients with visual hallucinations by psychosis type (i.e., either primary or secondary). Additional variables extracted were visual hallucination content, presence of non-visual hallucinations, diagnosis, study setting and patient characteristics. Variables were extracted using an adapted version of the Cochrane Collaboration data extraction form(10)

### Encoding Variables

We used the 10th revision of the International Statistical Classification of Diseases and Related Health Problems (ICD) to classify patients based on their initial diagnosis(11). Patients were classified as having a primary psychotic disorder if their psychotic symptoms were attributable to a primary non-affective (schizophrenia, schizotypal and delusional disorders; F20-F29) or affective psychotic disorder (F30-F39) following assessment and investigation. Patients were classified as having a secondary psychotic disorder if their psychotic symptoms were attributable to an organic condition, which included dementia (consistent with the category ‘Organic, including symptomatic, mental disorders’; F00-F09). Patients were also classified as having a secondary psychosis if their symptoms were attributable to medication or illicit substances (consistent with the category ‘mental and behavioural disorders due to psychoactive substance use’; F10-F19).

We also categorized the content of visual hallucinations using two overlapping constructs. First, visual hallucinations were classified based on complexity. We defined a complex visual hallucination as a formed object such as face, animal, or place and a simple hallucination by the absence of a formed object (e.g. flashing lights)(12). Second, visual hallucinations were classified based on whether they included people or animals (animate hallucinations) or inanimate objects, such as a vehicle or text (see supplementary table 5 for details of how hallucination content was classified).

### Risk of bias and quality assessment

Risk of bias was assessed using a modified version of the 8-item Newcastle-Ottawa Scale(13). Each study was scored out of 8 based on 7-items: case definition, case representativeness, selection of controls, definition of controls, comparability of cases and controls, ascertainment of cases and controls and non-response rate. Each study was independently rated by two researchers (A.K.D and M.T.D), with discrepancies resolved by a third researcher (G.B). Based on the total score, studies were categorised as being at high (≤ 3), medium (4-5) or low (6-7) risk of bias.

### Statistical analysis

Statistical analysis was conducted using R version 4.0.3 (R Core Team, Vienna, Austria; see https://cran.r-project.org) using the packages meta(14) and dmetar(15). The meta-analysis was performed using a random-effects model as methodological heterogeneity was anticipated. The DerSimonian and Laird inverse variance pooling method was used to calculate odds ratios (OR) and 95% confidence intervals (CI)(16). Findings were considered statistically significant if *p* < 0.05. Between-study heterogeneity was quantified using I^2^, categorised as low (25%), moderate (50%) or high (75%), and statistically assessed using chi-squared test (15,16). Meta regression was performed to explore the moderating effect of age. Subgroup analysis explored the moderating effect of type of secondary psychosis. This was implemented by stratifying studies based on the type of secondary psychosis included, namely: i) organic, ii) drug-induced (medication or illicit substance), or iii) organic *or* drug-induced psychosis.

Sensitivity analyses explored the impact of studies at high risk of bias. In addition, a leave-one-out sensitivity analysis was conducted to detect any influential studies, by iteratively removing one study at a time and recalculating the OR. Publication bias was evaluated through visual inspection of a funnel plot(17) and formally with Egger’s test(18).

Finally, secondary analyses were performed. First, we explored the association between visual hallucination content and psychosis type. We compared the groups in terms of the frequency of simple or complex visual hallucinations and separately in terms of frequency of animate or inanimate hallucinations, calculating the odds ratios for both.

Second, based on studies included as part of the primary meta-analysis, we explored the association between different hallucination modalities (auditory, tactile, olfactory, gustatory, or visual *and* auditory) and secondary psychosis, through calculation of odds ratios and a random-effects meta-analysis, following the same approach as described above.

## RESULTS

### Included studies

14 studies met eligibility and were published between 1965 and 2020(7,19–31). See PRISMA flow diagram for a summary of the selection process (supplementary figure 1)(32). Studies ranged in size between 14 and 200 patients and took place in Europe (N = 6), Asia (N = 4), Australia (N = 2), North America (N = 1) and unspecified (N=1). Characteristics of the included studies are reported (supplementary table 2).

### Meta-analysis

A total of 1,708 participants were included: 904 (52.9%) with primary psychosis and 804 (47.1%) with secondary psychosis. A random-effects model found a highly significant association between visual hallucinations and secondary psychosis with a pooled odds ratio of 3.0 (z = 4.0, 95% CI = 1.7-5.2, p < 0.001). There was moderate between study heterogeneity (I^2^ = 70%, χ^2^ (13) = 43.7, p *<* 0.001) (Figure 1).

**Fig 1.**
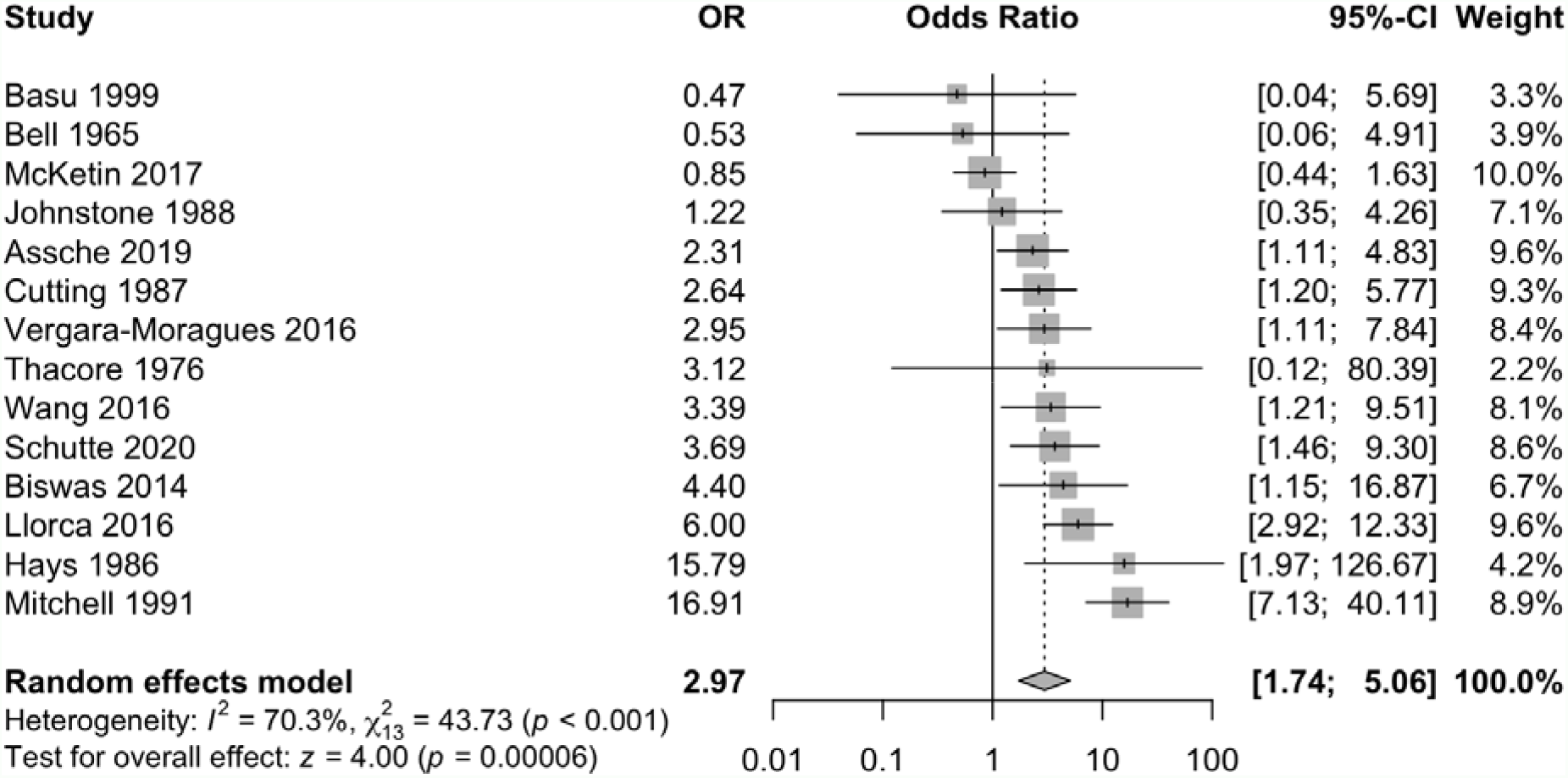
Forest plot displaying the random-effects meta-analysis results for the association between visual hallucinations and secondary psychosis

### Exploration of between study heterogeneity

Meta regression found age to be a non-significant moderator (QM (df = 1) = 0.33, *p* = 0.56) (supplementary figure 2). We grouped studies by the type of secondary psychosis reported: organic (OR = 3.73, 95% CI = 2.08-6.68), drug-induced (OR = 2.95, 95% CI = 1.16-7.50), and organic *or* drug-induced (OR = 2.10, 95% CI = 1.05-4.19). Subgroup analysis was non-significant (Q=1.55, *p* = 0.46) (supplementary figure 6).

### Risk of bias and publication bias assessment

Half the studies were rated as being at high risk of bias (n=7), with the remaining studies rated as medium (n=4) or low (n=3) risk (see supplementary table 3). The majority or studies did not report a history of disease (n=8). Most studies also did not report the non-response rate (n=10) or control for co-variables, such as age (n=10) (supplementary table 3). Funnel plot inspection (supplementary figure 4) indicated an absence of publication bias(17). This was supported by Egger’s test (*p* = 0.93).

### Sensitivity analyses

When removing studies at high risk of bias(21,23,26–30), the association between visual hallucinations and secondary psychosis increased (OR = 3.20, 95% CI = 1.8, 5.85, *z* = 3.90, p < 0.001) and heterogeneity reduced (I^2^ = 44%; χ ^2^ = 10.7, *p* = 0.10). Leave-one-out sensitivity analysis identified two influential studies (26,27) (supplementary figure 3). Notably, of these outlier studies, patients who experienced psychosis when using, *or* when abstaining from methamphetamine (“persistent MAP”) were categorised as secondary psychosis in the study by McKetin et al.(26). In addition, the study by Mitchell and Vierkant(27) was the only included study to have used a retrospective design. When McKetin et al.(26) was excluded, heterogeneity reduced to 55% (χ^2^ = 26.8, *p* < 0.001) and the OR increased to 3.5 (95% CI = 2.2 -5.6, *z* = 5.2, p < 0.001). When Mitchell and Vierkant (27) was excluded, heterogeneity reduced to 54% (χ^2^ = 26, *p* = 0.01) and the OR decreased to 2.5 (95% CI = 1.6 - 4.0, *z* = 4.0, p < 0.001). Finally, two post-hoc sensitivity analyses were performed: i) exclusion of studies where patients categorised as having a secondary cause of psychosis were later assigned a diagnosis of a primary psychosis (N=1)(21) and ii) exclusion of studies where visual hallucinations were aggregated with auditory hallucinations (N=1)(22). Significant and non-significant findings were unchanged under both sensitivity analysis.

### Secondary analyses

The content of visual hallucinations was reported in four studies(19,21,23,27) (See supplementary table 4). Information was available for 210 patients (51 primary psychosis patients; 142 secondary psychosis patients). Results are presented in Table 1. A significant association was found between inanimate visual hallucinations and secondary psychosis (OR = 0.1, 95% CI = 0.01 – 0.8, *p* = 0.03).

The presence of auditory, tactile, olfactory, gustatory, and visual *with* auditory hallucinations was reported in 14, 7, 5, 3 and 3 studies, respectively. Random-effects model found a significant association between olfactory hallucinations and secondary psychosis with a pooled odds ratio of 2.45 (95% CI = 1.09 - 5.52, *p* = 0.03). All other modalities were non-significant (see supplementary figure 5).

## DISCUSSION

This meta-analysis examined the association between visual hallucinations and secondary causes of psychosis in studies comparing primary and secondary psychosis directly. Across 14 studies, we had a pooled sample of 1,708 patients, consisting of 904 patients with a primary psychiatric disorder (predominantly schizophrenia and related disorders) and 804 with secondary psychosis. Our main finding was that patients with a secondary cause of psychosis were approximately three times more likely to experience visual hallucinations than patients with a primary psychiatric disorder. To our knowledge, this is the first study to investigate the association between visual hallucinations and psychosis aetiology in a meta-analysis of studies that have directly compared primary and secondary causes. Overall, our findings are consistent with the widely held view among clinicians that visual hallucinations are associated with secondary causes. Prior to this meta-analysis, however, this assertion was supported by observational studies of relatively modest sample sizes. Through pooling all published case-control studies, we were able to derive greater statistical precision.

### The content of visual hallucinations varies by psychosis aetiology

As a secondary objective, we explored whether there were differences in the content of visual hallucinations between primary and secondary causes of psychosis. Overall, irrespective of psychosis type, patients experiencing visual hallucinations were most likely to perceive a person or animal. However, those patients reporting inanimate objects, such as shadows or flashing lights, were more likely to have a secondary cause of psychosis.

### Between-study heterogeneity and robustness

There was moderate between-study heterogeneity which we explored using meta regression models and subgroup analyses. We did not find age, or type of secondary psychosis, to significantly change the findings. A leave-one-out sensitivity analysis revealed studies by McKetin et al.(26) and Mitchell and Vierkant(27) were outliers. When these studies were removed, the association between psychosis aetiology and visual hallucinations remained. Our findings also remained robust to removing studies at high risk of bias.

### Why are secondary causes of psychosis associated with visual hallucinations?

In this meta-analysis, the commonest cause of secondary psychosis was drug-induced psychosis, particularly cocaine, methamphetamine and cannabis use. We stratified studies by the type of patients with secondary psychosis included (organic, drug-induced, or either), however, we were unable to stratify by specific diagnosis (for example, Alzheimer’s disease and Parkinson’s disease). We did not find a significant difference between subgroups, suggesting patients with an organic *or* drug-induced cause of psychosis are comparable in their risk of developing visual hallucinations compared to primary psychosis.

Currently, our understanding on the mechanisms underpinning visual hallucinations is limited. It is not clear whether there are disease-dependent mechanisms(12) or a shared common neurophysiology across disorders(33) underpinning visual hallucinations(12,33). When viewed trans-diagnostically, there is evidence of brain changes associated with susceptibility to visual hallucinations(12) in visual networks, with occipital and parietal atrophy as a common neuroanatomical feature shared by primary and secondary causes of psychosis(33). Occipital atrophy is also found in patients with eye disease without psychosis who experience visual hallucinations (Charles Bonnet Syndrome), although it does not seem directly related to the neurophysiology of visual hallucinations as it is also found to the same degree in patients with eye disease who do not experience them(34). An emerging view is that the mechanism underlying visual hallucinations may be better captured by dynamic functional changes across visual networks, rather than structural brain changes alone(35). Such functional changes better explain some of the clinical contexts in which visual hallucinations are found, such as drug-induced psychosis or the early stages of neurodegenerative disease, where specific structural brain changes may not be apparent.

Why were patients with a secondary cause to their symptoms more likely to perceive an inanimate object? Animate objects (i.e. people and animals) are associated with the anterior ventral temporal lobe and para-hippocampal regions(36,37), whereas inanimate hallucinations are associated with posterior regions in the occipital lobe and surrounding areas(38). The implication is that secondary causes of psychosis are associated with more widespread cortical dysfunction involving occipital regions as well as anterior ventral temporal regions, perhaps reflecting the distribution of pathology in neurodegenerative disease or widely distributed effects in drug-induced causes. Although visual processing is altered in primary psychosis disorders such as schizophrenia(39), it is not thought these changes are central to the underlying mechanism of primary psychosis explaining why inanimate visual hallucinations may be infrequent.

As part of the secondary analyses, we also explored the association between non-visual hallucinations and secondary psychosis. We found olfactory hallucinations were the only other modality significantly associated with secondary psychosis. However, the effect size was lower than for visual hallucinations. These results suggest that across modalities, visual hallucinations *may* be the most strongly associated with secondary causes of psychosis, however, these results should be interpreted with caution (as we only included studies that met our eligibility criteria). Further research in this area is warranted.

### Clinical application

This study provides meta-analytic support for the association between secondary psychosis and visual hallucinations. As such, the findings provide the strongest evidence to date for routinely assessing patients for the presence of visual hallucinations as part of the standard psychiatric assessment. A positive finding may be considered a ‘red flag’ for a secondary cause. This quick and easy assessment may help clinicians in the calculus of deciding whether further investigation to exclude a secondary cause is indicated, for example through the use of neuroimaging (e.g. MRI) or neurophysiology (e.g. EEG). However, there are several caveats. As the psychopathology associated with psychosis is extremely broad, the absence of visual hallucinations should not be considered sufficient to *exclude* the possibility of an organic cause. Also, as the prevalence of visual hallucinations among different secondary causes of psychosis is unknown, it is difficult to determine how well the results from this meta-analysis generalise.

### Strengths

As the first meta-analysis exploring the association between secondary psychosis and visual hallucinations, this study provides the strongest evidence to date that visual hallucinations are associated with secondary causes of psychosis. A strength of restricting the inclusion of studies to those using a case-control design (as opposed to comparing the pooled proportion of visual hallucinations in different conditions) is that differences are more likely to be attributable to group, rather than study effects, as each study had broadly similar numbers of patients with primary and secondary cause of psychosis. A further strength of the study was the robustness of the findings to sensitivity analyses.

## Limitations

A limitation of restricting the inclusion of studies to case-control designs is that the number of studies included was restricted, reducing the overall statistical power. Secondary causes of psychosis are extremely diverse and there is likely to be a high degree of variability between specific causes and susceptibility toward visual hallucinations. This may explain some of the between-study heterogeneity, as studies varied in the type of secondary psychosis included. Due to the inherent nature of observational studies, causality between visual hallucinations and secondary psychosis could not be inferred. We were not able to adjust for potentially important confounding variables, such as medication, which may have inflated our effect size. For example, dopaminergic medication (commonly used to treat Parkinson’s disease) can induce visual hallucinations(40) and may have contributed to the large effect sizes observed in some studies(25). Finally, a high proportion of studies were rated being at a high risk of bias, however, we were partially able to evaluate the impact of this through a sensitivity analysis.

## Future research

Future research should focus on examining the association between visual hallucinations and subtypes of secondary psychosis. A more granular approach would be informative in terms of determining the specific associations between visual hallucinations and particular secondary causes of psychosis such as Parkinson’s disease, and Alzheimer’s disease. Additionally, research into the phenomenology of visual hallucinations is indicated to further delineate the nature of the association with secondary causes of psychosis. Finally, structural and functional neuroimaging studies are indicated to better understand the mechanisms underlying visual hallucinations in secondary psychosis which may help to identify potential novel treatment targets.

## Conclusion

Through a meta-analysis of case-control studies, visual hallucinations were found to be significantly associated with secondary causes of psychosis. Additionally, there was evidence that inanimate visual hallucinations are specifically associated with secondary psychosis. Results indicate that visual hallucinations may be considered by clinicians as a ‘red flag’ for a secondary psychosis. This may be particularly relevant for clinicians in deciding which patients warrant further investigation to exclude a secondary cause.

## Supporting information

references

## Data Availability

All data produced in the present study are available upon reasonable request to the authors

https://osf.io/j9twg/?view_only=5c0f63cccd574239aca07bccdae35f3c

## Declarations

### Statements and declarations

Preliminary findings were presented at the Faculty of Neuropsychiatry Annual Conference 2020 and at the British Neuropsychiatry Association Annual Meeting 2021.

## Acknowledgements

D.FF. and G.B. are supported by the NIHR South London and Maudsley NHS Foundation Trust Mental Health Biomedical Research Centre (BRC). D.FF. was supported by NIHR Programme Grants for Applied Research (RP-PG-0610-10100 - SHAPED). The views expressed are those of the authors and not necessarily those of the NIHR or the Department of Health and Social Care.

## Author contributions

A.K.D and G.B designed the study. A.K.D, M.T.D and G.B conducted the search and data extraction. A.K.D and G.B analysed the data and wrote the first draft of the manuscript. A.K.D, G.B, M.T.D and D.FF contributed and edited the manuscript.

### Data availability

The study was pre-registered and the data that support the findings of this study are available through the Open Science Framework (OSF) website (https://osf.io/j9twg/?view_only=5c0f63cccd574239aca07bccdae35f3c).

### Relevance statement

Visual hallucinations are associated with secondary causes of psychosis. Visual hallucinations of inanimate (i.e. no-living) objects, in particular, were associated with secondary causes. Findings suggest that the presence of visual hallucinations should be routinely assessed as part of a standard psychiatric assessment. A positive finding may be considered a ‘red flag’ for a secondary cause and warrant further investigation.

