## Supplementary materials for "The association between visual hallucinations and secondary psychosis: A systematic review and meta-analysis"

*For submission to Journal of Neurology, Neurosurgery and Psychiatry*

Supplementary materials

Supplementary Figure 1: PRISMA flow diagram…………………………………………………………….……………..……3

Supplementary Figure 2: Bubble plot displaying the meta-regression results…………………………..…..…..4

Supplementary Figure 3: Leave-one-out sensitivity analysis……………………………………………….……..………5

Supplementary Figure 4: Funnel plot of the meta-analysis studies………………………………………………..….5

Supplementary Figure 5: Forest plot displaying the random-effects meta-analysis

results for the association between various hallucination modalities and secondary psychosis……………………….……………………………………………………………………………………………………….….…6 - 8

Supplementary Figure 6: Forest plot displaying the subgroup analysis for

drug-induced psychosis………………………………………………………………………………………………………………..……9

Supplementary Table 1: PRISMA guidelines checklist…………………………………………………...……..…..10 - 15

Supplementary Table 2: Characteristics of studies included in the meta-analysis……………….…….16 - 23

Supplementary Table 3: Newcastle-Ottawa Scale scores for the assessment of

risk of bias……………………………………………………………………………………………………………………………………….24

Supplementary Table 4: Frequency of visual hallucinations experienced in

patients with primary and secondary psychosis…………………………………………………………………….……….25

Supplementary Table 5: Type of visual hallucinations experienced in patients with primary and

secondary psychosis……………………………………………………………………………………………………………..…………26

Supplementary Table 6: Studies excluded…………..…………………………………………………………..………….…..27

Supplementary Figure 1: PRISMA flow diagram

Records excluded
(n = 555)

Records screened
(n = 648)

Studies included in quantitative synthesis (meta-analysis)
(n = 14)

Full-text articles excluded, with reasons
(n = 79)

- No non-organic psychosis control group (n = 11)
- No organic psychosis (n = 24)
- No/insufficient information on visual hallucinations (n = 6)
- Non-English (n = 7)
- No access (n = 1)
- No full-text article available/conference abstract only (n = 11)
- Other (n = 19)

Full-text articles assessed for eligibility
(n = 93)

Records after duplicates removed
(n = 648)

### Identification

### Eligibility

### Included

### Screening

Additional records identified through other sources
(n = 6)

Records identified through database searching
(n = 885)

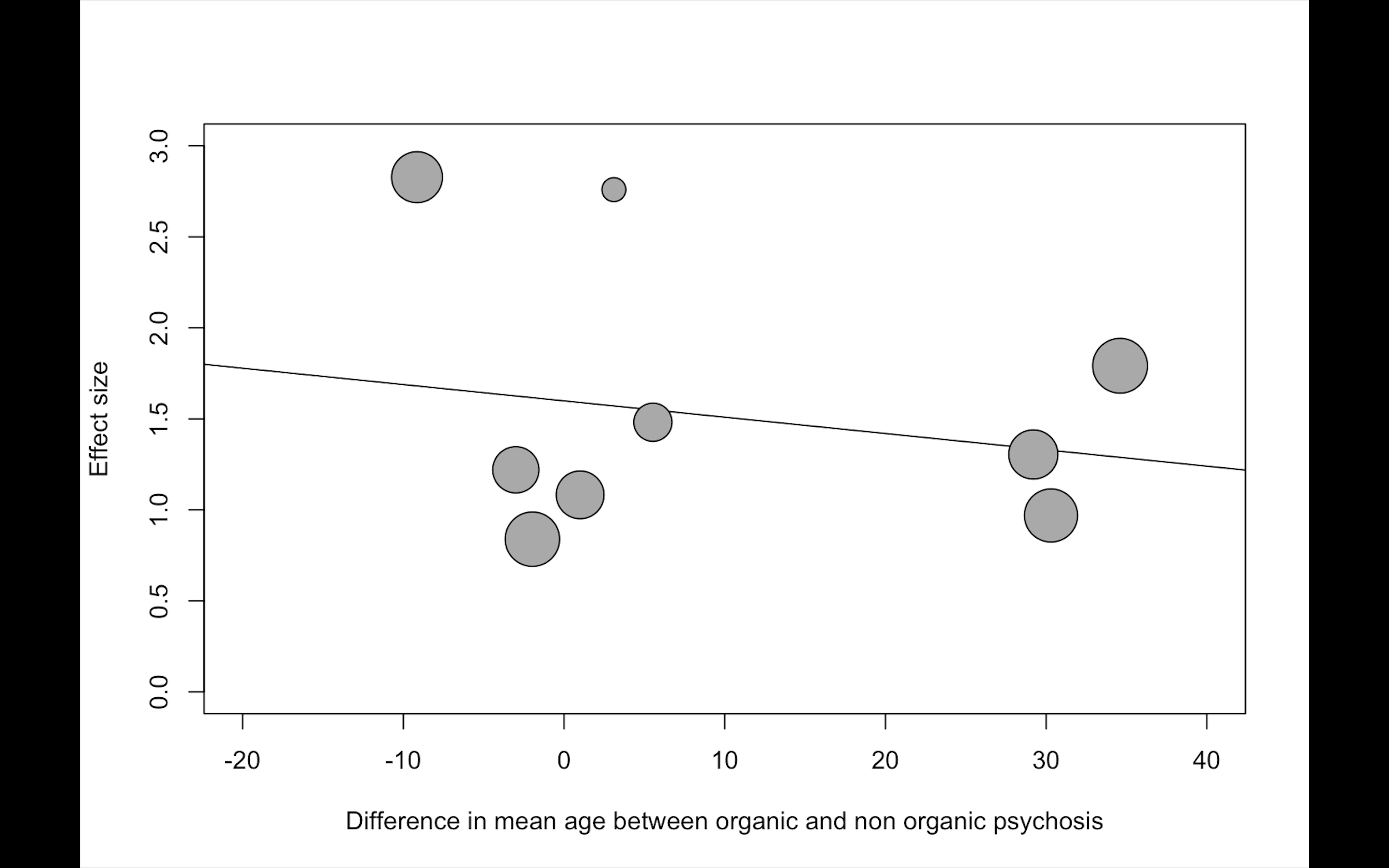

Supplementary Figure 2: Bubble plot displaying the meta-regression results.

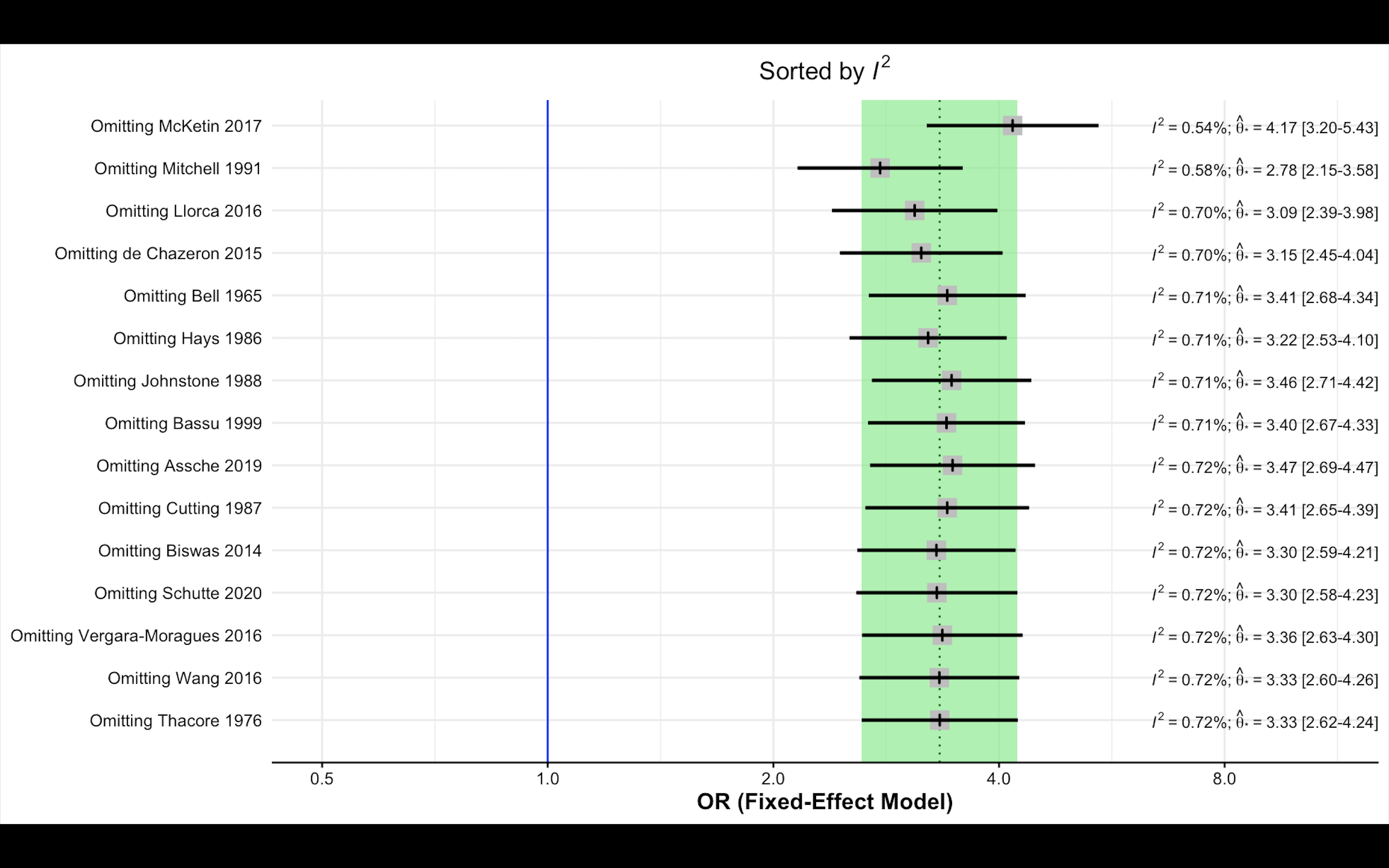

Supplementary Figure 3: Leave-one-out sensitivity analysis

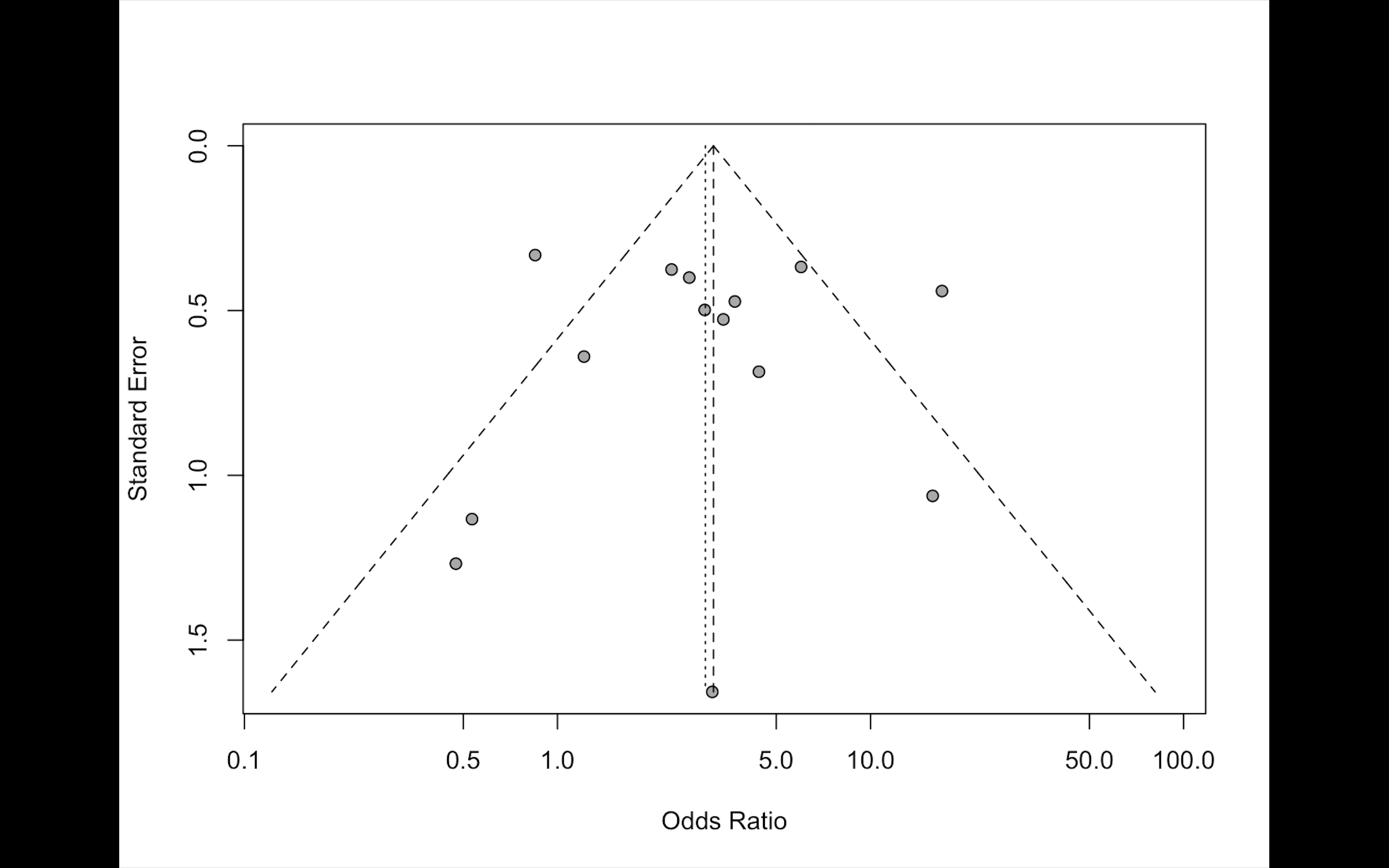

Supplementary Figure 4: Funnel plot of the meta-analysis studies.

a)

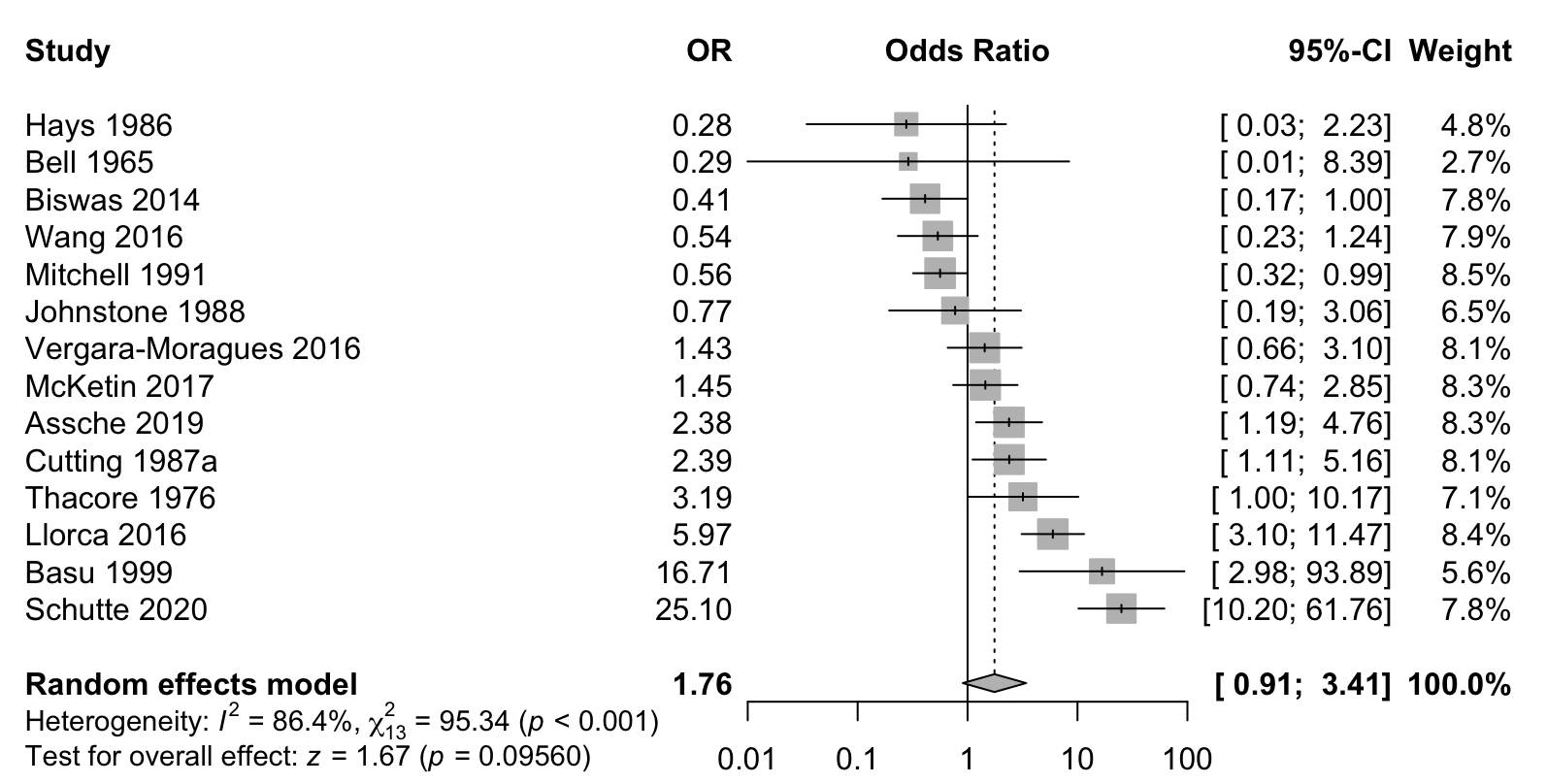

b)

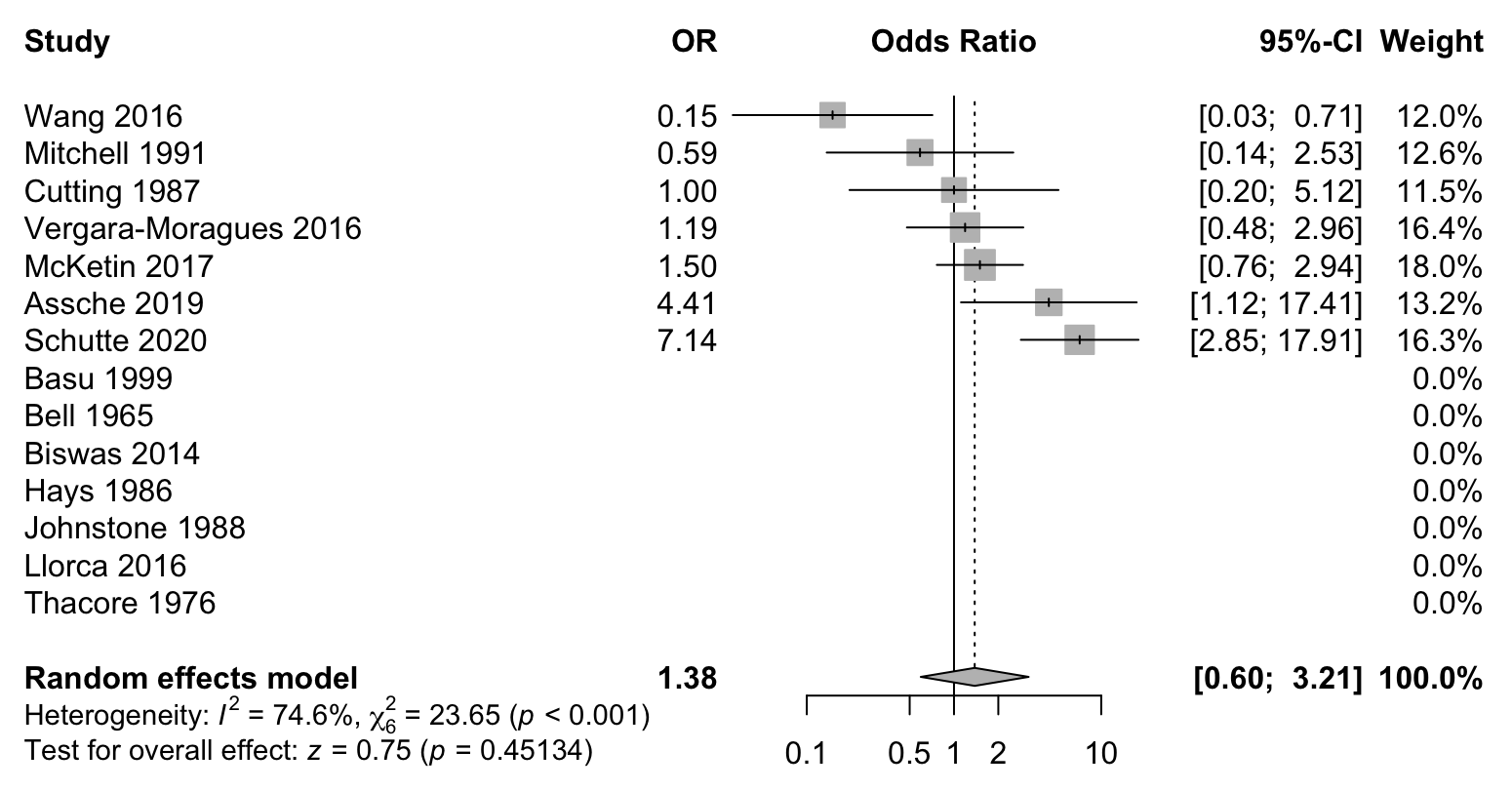

c)

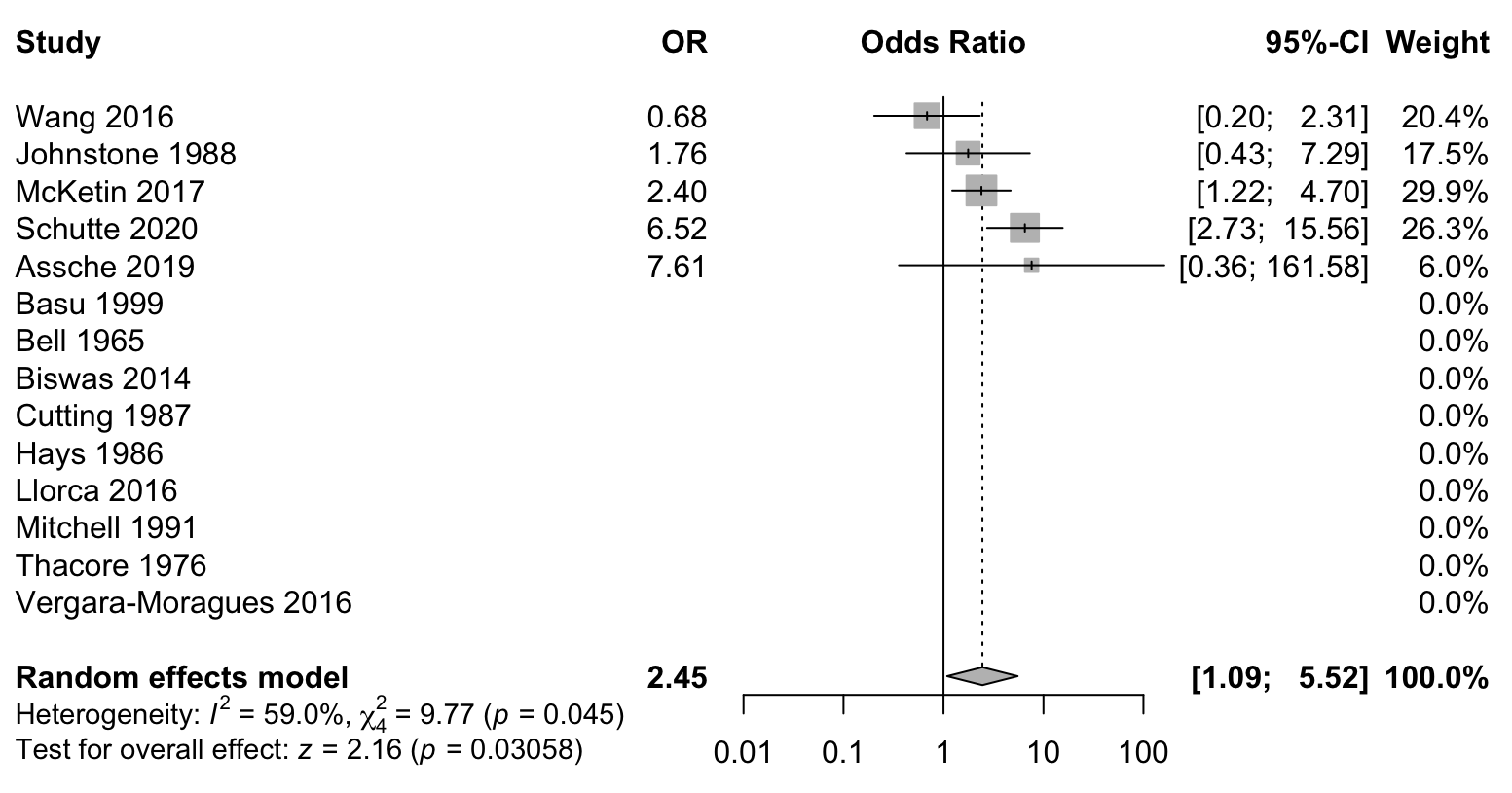

d)

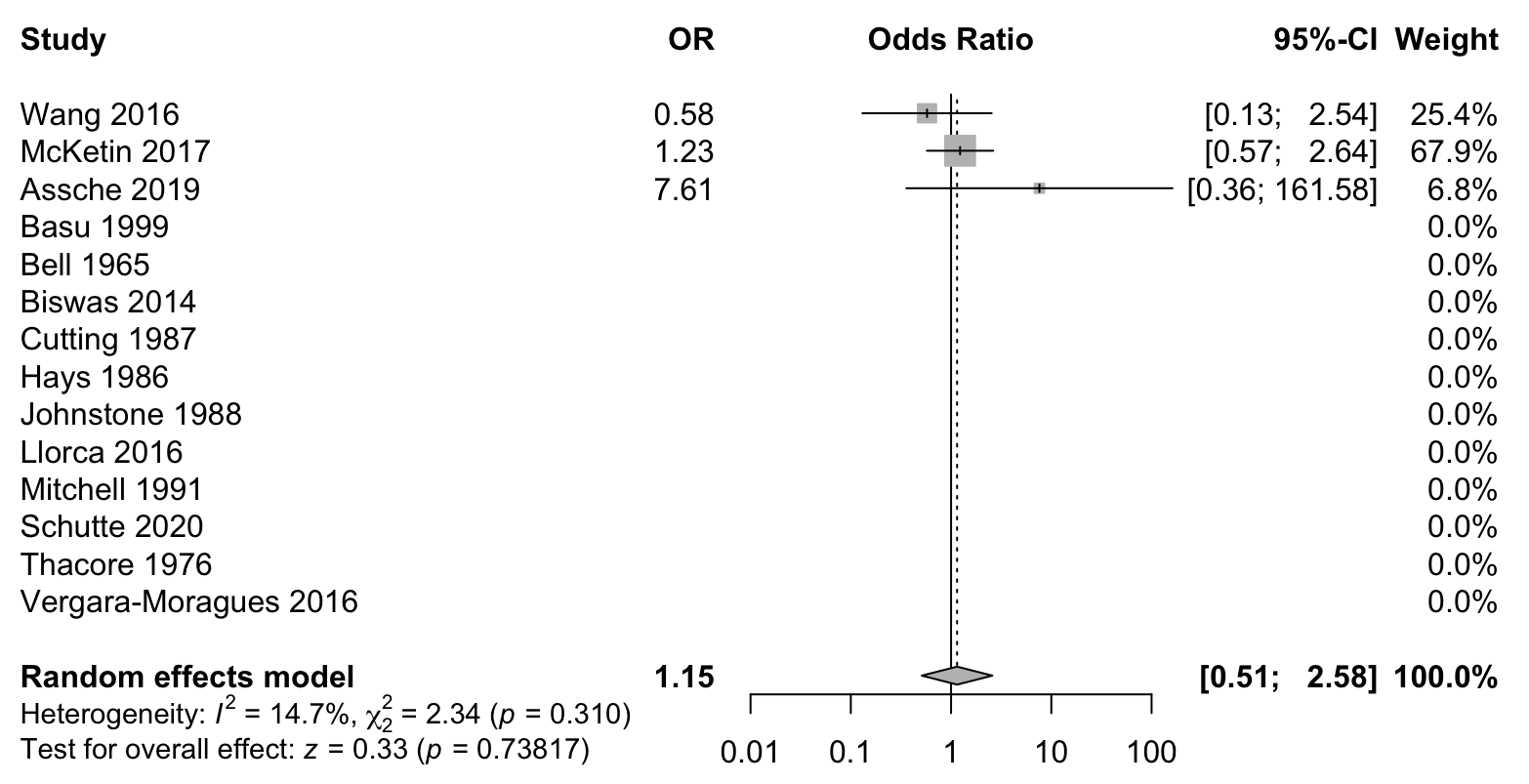

e)

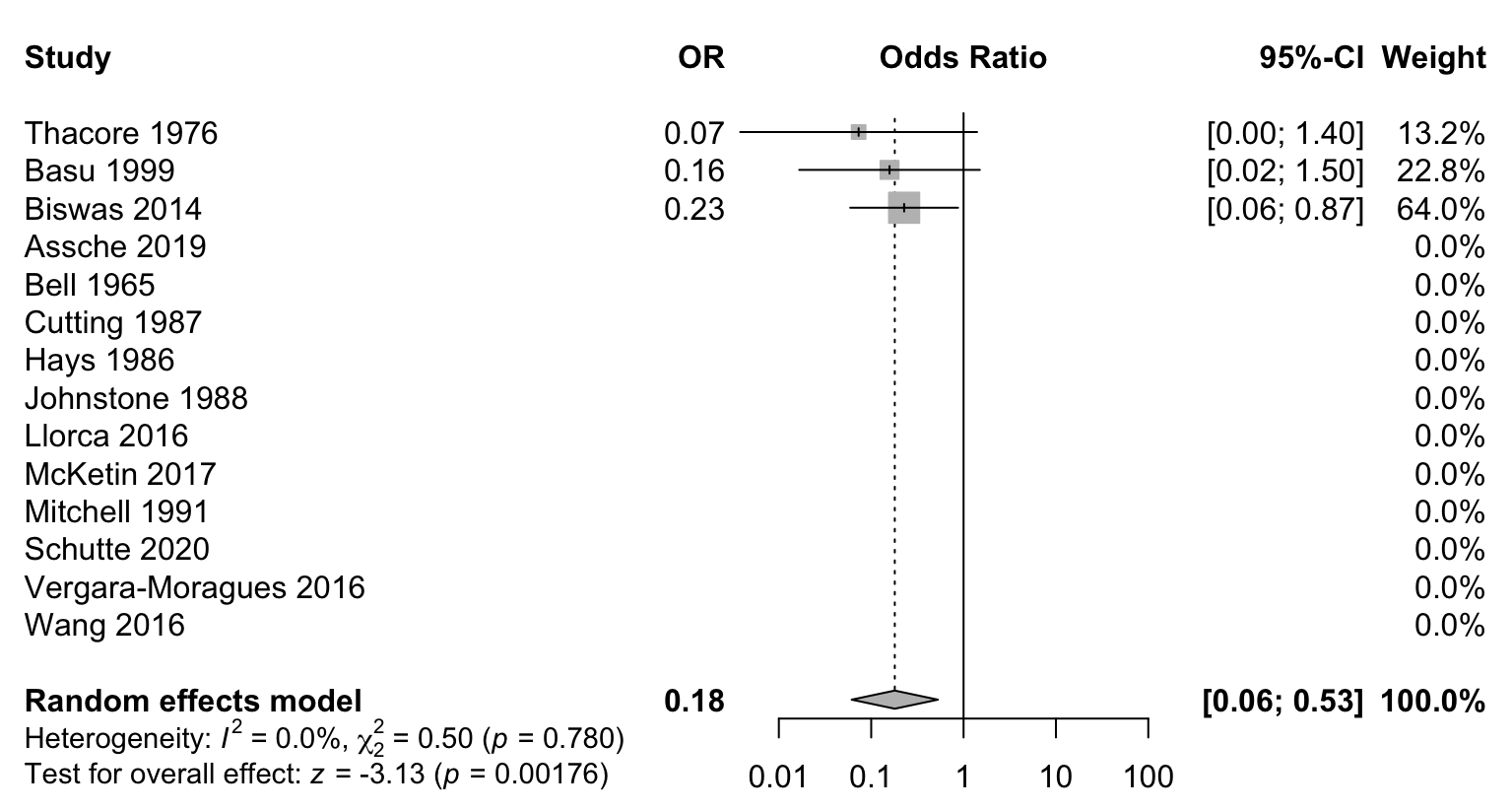

Supplementary Figure 5: Forest plots displaying the random-effects meta-analysis results for the association between secondary psychosis and a) auditory hallucinations, b) tactile hallucinations, c) olfactory hallucinations, d) gustatory hallucinations, e) visual and auditory hallucinations.

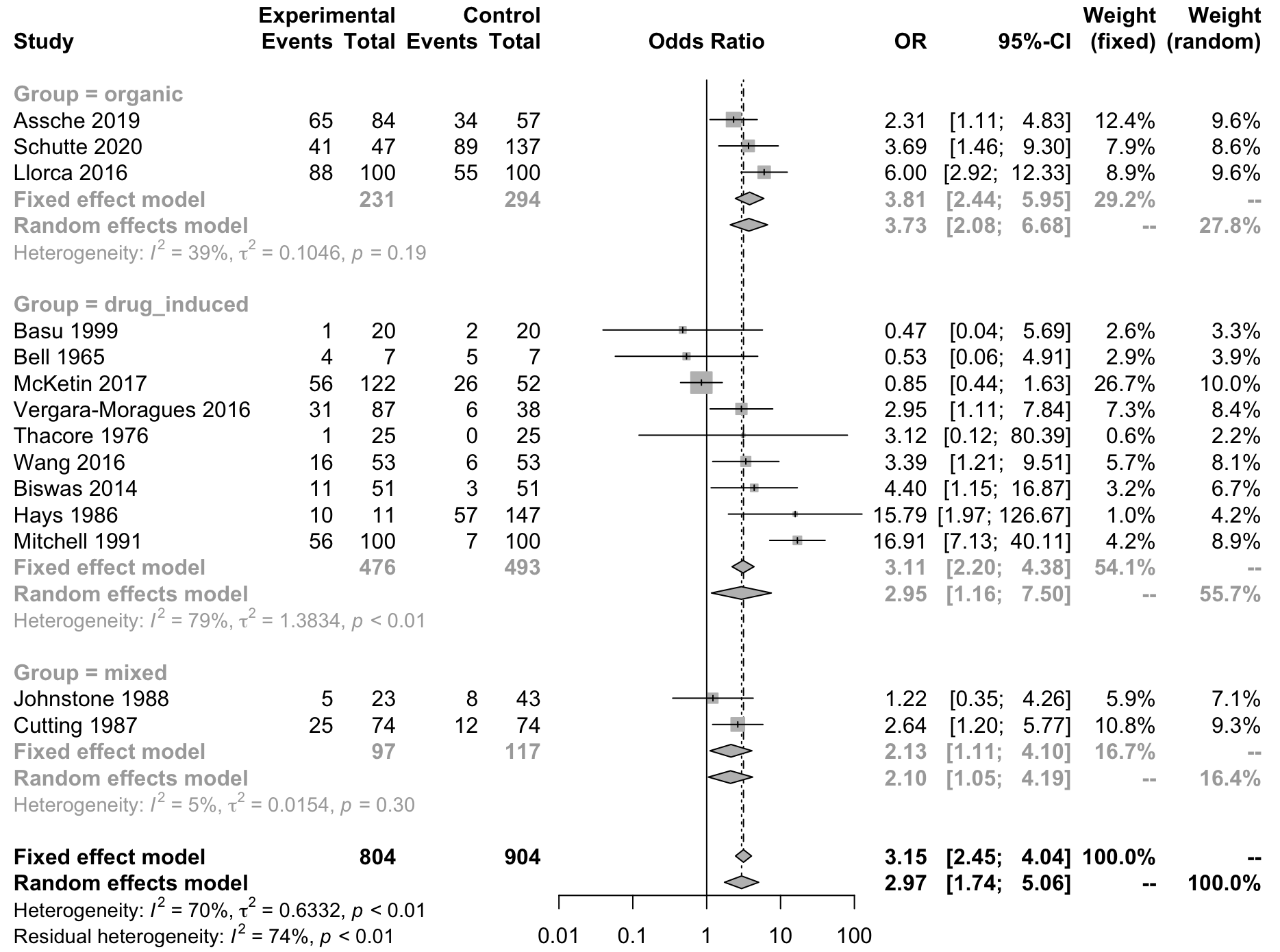

Supplementary Figure 6: Forest plot displaying the subgroup analysis for secondary psychosis a) organic, b) drug-induced and c) organic or drug-induced.

Supplementary table 1: PRISMA guidelines checklist

Table 1. PRISMA checklist

| **Section and Topic** | **Item #** | **Checklist item** | **Location where item is reported** |
| --- | --- | --- | --- |
| **TITLE** | | |  |
| Title | 1 | Identify the report as a systematic review. | 1 |
| **ABSTRACT** | | |  |
| Abstract | 2 | See the PRISMA 2020 for Abstracts checklist. | 2 |
| **INTRODUCTION** | | |  |
| Rationale | 3 | Describe the rationale for the review in the context of existing knowledge. | 3 |
| Objectives | 4 | Provide an explicit statement of the objective(s) or question(s) the review addresses. | 3 |
| **METHODS** | | |  |
| Eligibility criteria | 5 | Specify the inclusion and exclusion criteria for the review and how studies were grouped for the syntheses. | 4 |
| Information sources | 6 | Specify all databases, registers, websites, organisations, reference lists and other sources searched or consulted to identify studies. Specify the date when each source was last searched or consulted. | 4 |
| Search strategy | 7 | Present the full search strategies for all databases, registers and websites, including any filters and limits used. | 4 |
| Selection process | 8 | Specify the methods used to decide whether a study met the inclusion criteria of the review, including how many reviewers screened each record and each report retrieved, whether they worked independently, and if applicable, details of automation tools used in the process. | 4 |
| Data collection process | 9 | Specify the methods used to collect data from reports, including how many reviewers collected data from each report, whether they worked independently, any processes for obtaining or confirming data from study investigators, and if applicable, details of automation tools used in the process. | 4 |
| Data items | 10a | List and define all outcomes for which data were sought. Specify whether all results that were compatible with each outcome domain in each study were sought (e.g. for all measures, time points, analyses), and if not, the methods used to decide which results to collect. | 4 |
|  | 10b | List and define all other variables for which data were sought (e.g. participant and intervention characteristics, funding sources). Describe any assumptions made about any missing or unclear information. | 4 |
| Study risk of bias assessment | 11 | Specify the methods used to assess risk of bias in the included studies, including details of the tool(s) used, how many reviewers assessed each study and whether they worked independently, and if applicable, details of automation tools used in the process. | 4-5 |
| Effect measures | 12 | Specify for each outcome the effect measure(s) (e.g. risk ratio, mean difference) used in the synthesis or presentation of results. | 5 |
| Synthesis methods | 13a | Describe the processes used to decide which studies were eligible for each synthesis (e.g. tabulating the study intervention characteristics and comparing against the planned groups for each synthesis (item #5)). | N/A |
|  | 13b | Describe any methods required to prepare the data for presentation or synthesis, such as handling of missing summary statistics, or data conversions. | N/A |
|  | 13c | Describe any methods used to tabulate or visually display results of individual studies and syntheses. | 5-6 |
|  | 13d | Describe any methods used to synthesize results and provide a rationale for the choice(s). If meta-analysis was performed, describe the model(s), method(s) to identify the presence and extent of statistical heterogeneity, and software package(s) used. | 5-6 |
|  | 13e | Describe any methods used to explore possible causes of heterogeneity among study results (e.g. subgroup analysis, meta-regression). | 5-6 |
|  | 13f | Describe any sensitivity analyses conducted to assess robustness of the synthesized results. | 6 |
| Reporting bias assessment | 14 | Describe any methods used to assess risk of bias due to missing results in a synthesis (arising from reporting biases). | N/A |
| Certainty assessment | 15 | Describe any methods used to assess certainty (or confidence) in the body of evidence for an outcome. | 6 |
| **RESULTS** | | |  |
| Study selection | 16a | Describe the results of the search and selection process, from the number of records identified in the search to the number of studies included in the review, ideally using a flow diagram. | 7, Supplementary figure 1 |
|  | 16b | Cite studies that might appear to meet the inclusion criteria, but which were excluded, and explain why they were excluded. | Supplementary Table 6 |
| Study characteristics | 17 | Cite each included study and present its characteristics. | 7, Supplementary table 2 |
| Risk of bias in studies | 18 | Present assessments of risk of bias for each included study. | 8, Supplementary table 3 |
| Results of individual studies | 19 | For all outcomes, present, for each study: (a) summary statistics for each group (where appropriate) and (b) an effect estimate and its precision (e.g. confidence/credible interval), ideally using structured tables or plots. | 7, Fig 1 |
| Results of syntheses | 20a | For each synthesis, briefly summarise the characteristics and risk of bias among contributing studies. | 8, Supplementary table 3 |
|  | 20b | Present results of all statistical syntheses conducted. If meta-analysis was done, present for each the summary estimate and its precision (e.g. confidence/credible interval) and measures of statistical heterogeneity. If comparing groups, describe the direction of the effect. | 7, Fig 1 |
|  | 20c | Present results of all investigations of possible causes of heterogeneity among study results. | 8 |
|  | 20d | Present results of all sensitivity analyses conducted to assess the robustness of the synthesized results. | 8, Supplementary figure 3 |
| Reporting biases | 21 | Present assessments of risk of bias due to missing results (arising from reporting biases) for each synthesis assessed. | N/A |
| Certainty of evidence | 22 | Present assessments of certainty (or confidence) in the body of evidence for each outcome assessed. | Supplementary Figure 4 |
| **DISCUSSION** | | |  |
| Discussion | 23a | Provide a general interpretation of the results in the context of other evidence. | 10 |
|  | 23b | Discuss any limitations of the evidence included in the review. | 12-13 |
|  | 23c | Discuss any limitations of the review processes used. | 12-13 |
|  | 23d | Discuss implications of the results for practice, policy, and future research. | 12 |
| **OTHER INFORMATION** | | |  |
| Registration and protocol | 24a | Provide registration information for the review, including register name and registration number, or state that the review was not registered. | 14 |
|  | 24b | Indicate where the review protocol can be accessed, or state that a protocol was not prepared. | 14 |
|  | 24c | Describe and explain any amendments to information provided at registration or in the protocol. | N/A |
| Support | 25 | Describe sources of financial or non-financial support for the review, and the role of the funders or sponsors in the review. | 14 |
| Competing interests | 26 | Declare any competing interests of review authors. | 14 |
| Availability of data, code and other materials | 27 | Report which of the following are publicly available and where they can be found: template data collection forms; data extracted from included studies; data used for all analyses; analytic code; any other materials used in the review. | 14 |

Supplementary table 2: Characteristics of studies included in the meta-analysis.

| Study (author, year) | Type of psychosis | Sample (n) | Visual hallucinations (%) | Age (years) | Gender (% males) | Setting and country |
| --- | --- | --- | --- | --- | --- | --- |
| Assche et al., 2019 | Secondary: DLB + AD  Primary: Very-late-onset schizophrenia-like psychosis | Secondary: 84 (DLB: 49;AD: 35)  Primary: 57 | Secondary: 65/84 (77)  Primary: 34/57 (60) | DLB: 76.2 (mean)  AD: 78.8 (mean)  Primary: 79.25 (mean) | DLB: 67.3  AD: 37.1  Primary: 22.8 | Secondary: General hospital, Belgium  Primary: General hospital, Belgium |
| Basu et al., 1999 | Secondary: Cannabis psychosis  Primary: SCZ | Secondary: 20  Primary: 20 | Secondary: 1/20 (5)  Primary: 2/20 (10) | N/A (age-matched) | 100 | Secondary: Drug Addiction and Treatment Centre, India  Primary: General psychiatry register, India |
| Bell, 1965 | Secondary: Amphetamine psychosis  Primary: SCZ | Secondary: 7  Primary: 7 | Secondary: 4/7 (57)  Primary: 5 (71) | N/A | N/A | Secondary: Psychiatric hospital, Australia  Primary: Psychiatric hospital, Australia |
| Biswas et al., 2014 | Secondary: Chloroquine psychosis  Primary: Brief psychotic disorder | Secondary: 51  Primary: 51 | Secondary: 11/51 (22)  Primary: 3/51 (6) | Secondary: 31.29 (mean)  Primary: 25.76 (mean) | Secondary: 52.9  NOP: 47.1 | Secondary: Psychiatric hospital, India  Primary: Psychiatric hospital, India |
| Cutting, 1987 | Secondary: Alcohol, respiratory failure, carcinoma, liver failure, myxoedema, right parietal cerebrovascular accident, drugs, renal failure, cardiac failure, Systemic lupus erythematosus  Primary: Acute SCZ | Secondary: 74  Primary: 74 | Secondary: 25/74 (33)  Primary: 12/74 (16) | Secondary: 57.4 (mean)  Primary: 27.1 (mean) | Secondary: 50.0  Primary: 55.4 | Secondary: Psychiatric referrals from the general wards of two hospitals, England  Primary: acute admission ward of a psychiatric hospital |
| Hays & Aidroos, 1986 | Secondary: Alcohol psychosis  Primary: SCZ | Secondary: 11  Primary: 147 | Secondary: 10/11 (91)  Primary: 57/147 (39) | Secondary: 28.8 (mean)  Primary: 25.7 (mean) | Secondary: 81.8  Primary: N/A | N/A |
| Johnstone et al., 1988 | Secondary: Drug abuse/withdrawal (amphetamine, alcohol, heroin, cannabis, barbituates, diazepam, ephedrine) + hypothyroidism, thyrotoxicosis, syphyllis, cerebrovascular accident, cancer (brain, lung), ulcerative colitis on steroids, Systemic lupus erythematosus, B_12_ deficiency, uncontrolled type 1 diabetes  Primary: N/A | Secondary: 23  Primary: 43 | Secondary: 5/23 (22)  Primary: 8/43 (19) | Secondary: 40.7 (mean) (matched perfectly to 16 Primary: controls) | Secondary: 56.5 (matched perfectly to 16 Primary: controls) | Secondary: General hospital, England  Primary: General hospital, England |
| Llorca et al., 2016 | Secondary: Parkinson’s disease  Primary: SCZ | Secondary: 100  Primary: 100 | Secondary: 88/100 (88)  Primary: 55/100 (55) | Secondary: 71.1 (mean)  Primary: 36.5 (mean) | Secondary: 55  Primary: 69 | Five psychiatric departments and two neurological departments, France |
| McKetin et al., 2017 | Secondary: Methamphatamine psychosis  Primary: N/A | Secondary: 122  Primary: 52 | Secondary: 56/122 (46)  Primary: 26/52 (50) | 31.6 (mean) | 71 | Secondary: Drug Addiction and Treatment Centre, Australia^1^  Primary: Recruited from the Methamphetamine Treatment Evaluation Study (MATES), Australia^1^ |
| Mitchell & Vierkant, 1991 | Secondary: Cocaine psychosis  Primary: Paranoid SCZ | Secondary: 100  Primary: 100 | Secondary: 56/100 (56)  Primary: 7/100 (7) | Secondary: 28.58 (mean)  Primary: 34.73 (mean) | Secondary: 71  Primary: 56 | Secondary: Psychiatric hospital, Texas  Primary: Psychiatric hospital, Texas |
| Schutte et al., 2020 | Secondary: DLB + PD + AD  Primary: SCZ + mood disorder + borderline personality disorder + post-traumatic stress disorder | Secondary: 47  Primary: 137 | Secondary: 41/47 (87)  Primary: 89/137 (65) | Secondary: 43.6 (mean)  Primary: 39.9 (mean) | Secondary: 13.6  Primary: 55.1 | Psychiatry and neurology departments of a general hospital and psychiatric hospital, Netherlands |
| Thacore & Shukla, 1976 | Secondary: Cannabis psychosis  Primary: Paranoid SCZ | Secondary: 25  Primary: 25 | Secondary: 1/25 (4)  Primary: 0 (0) | Secondary: 18-42 (range)  Primary: 16-50 (range) | 100 | N/A, India |
| Vergara-Moragues et al., 2016 | Secondary: Cocaine psychosis  Primary: Independent psychotic disorder | Secondary: 87  Primary: 38 | Secondary: 31/87 (36)  Primary: 6/38 (16) | Secondary: 36 (mean)  Primary: 35 (mean) | Secondary: 8  Primary: 30 | Drug Addiction and Treatment Centres, outpatient drug dependence programme, detoxification unit of a general hospital, targeted sampling in street sites^2^, Spain |
| Wang et al., 2016 | Secondary: Methamphatamine psychosis  Primary: SCZ | Secondary: 53  Primary: 53 | Secondary: 16/53 (30)  Primary: 6/53 (11) | Secondary: 36.8 (mean)  Primary: 39.8 (mean) | Secondary: 77.4  Primary: 67.9 | Secondary: Two general hospitals and two psychiatric hospitals, Taiwan  Primary: General hospital, Taiwan |

Note. DLB = Dementia with Lewy Bodies. AD = Alzheimer’s disease. PD = Parkinson’s Disease. SCZ = schizophrenia. N/A = not available. ^1^Participants were selected from a larger study, the Methamphetamine Treatment Evaluation Study (MATES), Australia(41). ^2^Participants were recruited from a larger study(42).

Supplementary table 3: Newcastle-Ottawa Scale scores for the assessment of risk of bias. S = Selection, C = Comparability, E = Exposure.

| Study | S1 | S2 | S3 | S4 | C1 | E1 | E2 | E3 | Total | Risk |
| --- | --- | --- | --- | --- | --- | --- | --- | --- | --- | --- |
| Assche 2019 | ★ | ★ |  |  | ★★ |  | ★ |  | 5 | Medium |
| Basu 1999 | ★ | ★ |  | ★ | ★★ |  | ★ | ★ | 7 | Low |
| Bell 1965 |  |  |  |  |  |  | ★ |  | 1 | High |
| Biswas 2014 | ★ | ★ |  | ★ | ★★ |  | ★ |  | 6 | Low |
| Cutting 1987 | ★ | ★ |  |  | ★ |  |  |  | 3 | High |
| Hays 1986 | ★ | ★ |  | ★ |  |  | ★ |  | 4 | Medium |
| Johnstone 1988 | ★ | ★ |  |  | ★★ |  | ★ | ★ | 6 | Medium |
| Llorca 2016 | ★ | ★ |  | ★ |  |  | ★ |  | 4 | Medium |
| McKetin 2017 | ★ | ★ |  |  |  |  | ★ | ★ | 4 | High |
| Mitchell 1991 | ★ | ★ |  |  |  |  | ★ |  | 3 | High |
| Schutte 2020 | ★ | ★ |  |  |  |  | ★ |  | 3 | High |
| Thacore 1976 |  |  |  |  | ★★ |  | ★ | ★ | 4 | High |
| Vergara-Moragues 2016 |  | ★ |  | ★ |  |  | ★ |  | 3 | High |
| Wang 2016 | ★ | ★ |  | ★ | ★★ |  | ★ |  | 6 | Low |

Supplementary Table 4. Frequency of visual hallucinations experienced in patients with primary and secondary psychosis. Patients may have multiple types of visual hallucinations.

| Visual hallucination | Primary (N=52) | Secondary (N=158) |
| --- | --- | --- |
|  | N (%) | N (%) |
| People and/or animals | 48 (92.3) | 117 (74.1) |
| Unspecified object | 1 (1.92) | 16 (10.1) |
| Shadows | 0 (0) | 10 (6.33) |
| Flashing lights | 0 (0) | 9 (5.70) |
| Vehicles | 0 (0) | 2 (1.27) |
| Other | 3 (5.77) | 4 (7.69) |

Note. ‘Unspecified objects’ was excluded from the secondary analysis

Supplementary Table 5. Type of visual hallucinations experienced in patients with primary and secondary psychosis. Patients may have multiple types of visual hallucinations.

| Visual hallucination | Primary (N=52) | | | | Secondary (N=158) | | | |
| --- | --- | --- | --- | --- | --- | --- | --- | --- |
|  | Animate | Inanimate | Simple | Complex | Animate | Inanimate | Simple | Complex |
| People and/or animals | X |  |  | X | X |  |  | X |
| Unspecified objects |  | X |  | X |  | X |  | X |
| Shadows |  | X |  | X |  | X |  | X |
| Flashing lights |  | X | X |  |  | X | X |  |
| Vehicles |  | X |  | X |  | X |  | X |
| Blurred visions^1^ |  | X |  | X |  | X |  | X |
| God and demons | X |  |  | X | X |  |  | X |
| Mirage |  | X |  | X |  | X |  | X |
| Witches, goblins, gremlins | X |  |  | X | X |  |  | X |
| Words |  | X |  | X |  | X |  | X |

Note. ^1^ “Blurs of visions of things”

Supplementary Table 6. studies excluded

| Reference | Reason for exclusion |
| --- | --- |
| de Chazeron, I., Pereira, B., Chereau-Boudet, I., Brousse, G., Misdrahi, D., & Fénelon, G. et al. (2015). Validation of a Psycho-Sensory hAllucinations Scale (PSAS) in schizophrenia and Parkinson's disease. *Schizophrenia Research*, *161*(2-3), 269-276. https://doi.org/10.1016/j.schres.2014.11.010 | Same cohort used in the study ‘Hallucinations in schizophrenia and Parkinson’s disease: an analysis of sensory modalities involved and the repercussion on patients’ (Llorca et al., 2016) |
| Lowe, G. (1973). The Phenomenology of Hallucinations as an Aid to Differential Diagnosis. *British Journal Of Psychiatry*, *123*(577), 621-633. https://doi.org/10.1192/bjp.123.6.621 | Insufficient data reported |
| Rottanburg, D., Robins, A., Ben-Arie, O., Teggin, A., & Elk, R. (1982). Cannabis-Associated Psychosis With Hypomanic Features. *The Lancet*, *320*(8312), 1364-1366. https://doi.org/10.1016/s0140-6736(82)91270-3 | Insufficient data reported |
| Tsuang, M., Simpson, J., & Kronfol, Z. (1982). Subtypes of Drug Abuse With Psychosis. *Archives Of General Psychiatry*, *39*(2), 141. https://doi.org/10.1001/archpsyc.1982.04290020013003 | Insufficient data reported |
