## Supplementary material for "The association between visual hallucinations and secondary psychosis: A systematic review and meta-analysis": references

10.	Cochrane Collaboration. Effective Practice and Organisation of Care (EPOC). Data collection form. EPOC Resources for review authors. Oslo: Norwegian Knowledge Centre for the Health Services; 2013. . 2013.

11.	World Health Organization(WHO). The ICD-10 classification of mental and behavioural disorders. 1993;

12.	O’Brien J, Taylor JP, Ballard C, Barker RA, Bradley C, Burns A, et al. Visual hallucinations in neurological and ophthalmological disease: pathophysiology and management. Journal of Neurology, Neurosurgery & Psychiatry. 2020 May;91(5):512–9.

13.	Wells GA, Shea B, O’Connell D, Peterson J, Welch V, Losos M, et al. The Newcastle-Ottawa Scale (NOS) for assessing the quality of nonrandomised studies in meta- analyses. 2013.

14.	Balduzzi S, Rücker G, Schwarzer G. How to perform a meta-analysis with R: a practical tutorial. Evidence Based Mental Health. 2019 Nov;22(4):153–60.

15.	Harrer M, Cuijpers P, Furukawa T, Ebert DD. dmetar: Companion R Package For The Guide “Doing Meta-Analysis in R”. R package version 0.0.9000. 2019.

23.	Cutting J. The Phenomenology of Acute Organic Psychosis Comparison with Acute Schizophrenia. British Journal of Psychiatry. 1987;151.

32.	Moher D, Liberati A, Tetzlaff J, Altman DG. Preferred reporting items for systematic reviews and meta-analyses: the PRISMA statement. BMJ. 2009 Jul 21;339(jul21 1):b2535–b2535.
